# Tandem Mass Tag-Based Serum Proteomics Reveals Distinct Apolipoprotein Signatures in Diabetic and Coronary Artery Disease Patients

**DOI:** 10.64898/2025.12.01.25341339

**Authors:** Abinayaa Rajkumar, Anil Kumar Challa, Nang Zhang, Krishna Parsawar, Gokul Panchalingam, M Suresh, Rajasekaran Namakkal-Soorappan, Kalaiselvi Periandavan, Sini Sunny

**Affiliations:** Department of Medical Biochemistry, University of Madras, Chennai, Tamil Nadu, India; Cardiac Aging & Redox Signaling Laboratory, Molecular and Cellular Pathology, Department of Pathology/Center for Free Radical Biology, University of Alabama at Birmingham, Birmingham, AL, USA; Analytical and Biological Mass Spectrometry Core Facility, University of Arizona, Tuscon, USA; Global Gleneagles Hospitals, Chennai, Tamil Nadu, India

**Keywords:** TMT proteomics, apolipoprotein oxidation, diabetes, coronary artery disease, redox biomarkers, lipid metabolism

## Abstract

Coronary artery disease (CAD) and diabetes mellitus (DM) frequently coexist, accelerating atherosclerotic progression through oxidative and metabolic dysregulation. However, the distinct serum proteomic signatures that differentiate CAD alone from CAD with DM comorbidity remain poorly characterized. We employed tandem mass tag (TMT)–based quantitative proteomics to delineate the molecular and oxidative alterations that distinguish CAD alone from CAD with diabetic comorbidity, with an emphasis on apolipoprotein remodeling and redox-lipid interactions. Serum samples from healthy controls, CAD, and diabetic cohorts (n = 6/group) were subjected to high-resolution LC–MS/MS analysis and redox biomarker profiling (LPO, GSH, PON1). Differentially expressed proteins and oxidative post-translational modifications (PTMs) were analyzed using gene ontology and protein–protein interaction (PPI) network approaches. TMT proteomics identified distinct redox–metabolic signatures segregating the three cohorts. CAD serum exhibited enrichment of inflammatory, proteolytic, and extracellular matrix remodeling pathways, whereas diabetic samples showed dominant metabolic and ER-stress networks. Notably, cysteine oxidation in ApoE was unique to diabetic patients, while lysine oxidation in ApoB and cysteine oxidation in ApoD characterized CAD, revealing disease-specific oxidative remodeling of apolipoproteins. PPI network mapping positioned ApoB and ApoE as central hubs linking lipid transport, coagulation, and complement cascades. This study provides the first redox-informed serum proteomic atlas distinguishing diabetic and CAD patients, uncovering oxidative PTMs in apolipoproteins as key determinants of lipoprotein dysfunction and atherogenic risk.

## Introduction

Atherosclerotic coronary artery disease (CAD) remains a leading cause of cardiovascular morbidity and mortality, and its progression is markedly accelerated in the presence of diabetes mellitus (DM) [1–4]. Despite extensive clinical research on CAD [5–8] and diabetes [9–12] as independent entities, the intersecting molecular mechanisms through which diabetic comorbidity exacerbate coronary pathology remain incompletely defined. Clinical and experimental studies indicate that diabetes profoundly alters lipid metabolism, oxidative homeostasis, and inflammatory signaling [13–16], yet the composite serum proteomic and oxidative signatures that distinguish CAD alone from CAD with diabetic complications have not been comprehensively elucidated.

Previous serum proteomic studies have largely examined CAD and diabetic cohorts separately [5–12], overlooking the synergistic metabolic and redox perturbations that emerge when both conditions coexist. In particular, how diabetes-associated comorbidity reprograms the circulating proteome in CAD patients, potentially amplifying lipoprotein dysfunction, endothelial injury, and plaque instability remains poorly understood. Given that diabetic CAD represents a major clinical phenotype characterized by heightened oxidative and carbonyl stress [17–19], there is a critical need to identify proteome-level modifications that uniquely define this comorbid state.

Apolipoproteins are central regulators of lipid transport, reverse cholesterol efflux, and antioxidant defense in plasma [20–22]. Alterations in apolipoprotein composition and function have been consistently linked to both dyslipidemia and atherogenesis [23, 24]. However, the site-specific oxidative modifications of apolipoproteins including methionine sulfoxidation, cysteine S-thiolation, tryptophan oxidation, and lysine carbonylation have not been systematically mapped in the context of CAD with diabetic complications. Such oxidative alterations can profoundly affect lipoprotein conformation, receptor binding, and HDL functionality [25–27], thereby shifting the serum proteome toward a more atherogenic and pro-inflammatory state.

To address these gaps, we employed tandem mass tag (TMT)-based quantitative serum proteomics to dissect the molecular differences between CAD alone and CAD with diabetic comorbidity. This unbiased approach enables simultaneous quantitation of hundreds of circulating proteins and detection of oxidative and post-translational modifications (PTMs) at the amino acid level [28–30]. By integrating quantitative redox proteomics with pathway enrichment and apolipoprotein-centric analysis, we aimed to delineate how diabetic comorbidity amplifies lipid-transport abnormalities in CAD.

Our study provides the first comprehensive apolipoprotein proteomic signature of CAD with diabetic complications, revealing specific oxidative modifications on apolipoproteins that may underlie dysfunctional lipid handling and accelerated atherogenesis. These findings not only offer mechanistic insights into redox dysregulation at the lipid–protein interface but also identify potential redox biomarkers and therapeutic targets relevant to cardiometabolic disease progression.

## Methods

### Sample Collection, Processing and Ethical considerations

Blood samples were collected from apparently healthy individuals attending the Master Health Checkup clinics at Global Gleneagles Hospitals and Sri Ramachandra Medical College (SRMC) Hospitals, Chennai, India. Fasting blood samples were also obtained from patients attending the cardiology and diabetology wards of the same institutions (n = 6 per group), including individuals with type 2 diabetes and those diagnosed with coronary artery disease (CAD). Approximately 5–10 mL of venous blood was drawn into EDTA-coated tubes and centrifuged at 3,000 × g for 10 minutes at 4°C to separate plasma and serum fractions [31]. All samples were aliquoted and stored at –80°C until biochemical and proteomic analyses were performed. All participants provided written informed consent prior to sample collection. The study was conducted in accordance with the Declaration of Helsinki and approved by the institutional ethics committees of all participating institutions (University of Madras: UM/IHEC/F.RM/2021-XIV; Global Gleneagles Hospitals: BMHR/2024/0076; SRMC: IEC-NI/21/APR/78/85).

### Sample preparation for Tandem Mass Tag (TMT) proteomic analysis

Briefly, serum protein concentrations were measured using a BCA kit [32] (ThermoFisher Scientific, Waltham, MA, USA) and 50 ug of each protein mixture was digested with trypsin at 47°c for 2hrs. Protein digestion by S-trap was performed by following the manufactory protocol [33]. The peptides were labeled using TMT 10plex reagents [34, 35]. The reagents were aliquoted into two groups for a total of 18 samples. Two labels were used as a pooled sample for each group. The reactions were stopped by adding hydroxylamine to a final concentration of 0.4%. Equal peptide amounts from each sample were combined for LC-MS/MS analysis (**Figure 1**).

**Figure 1.**
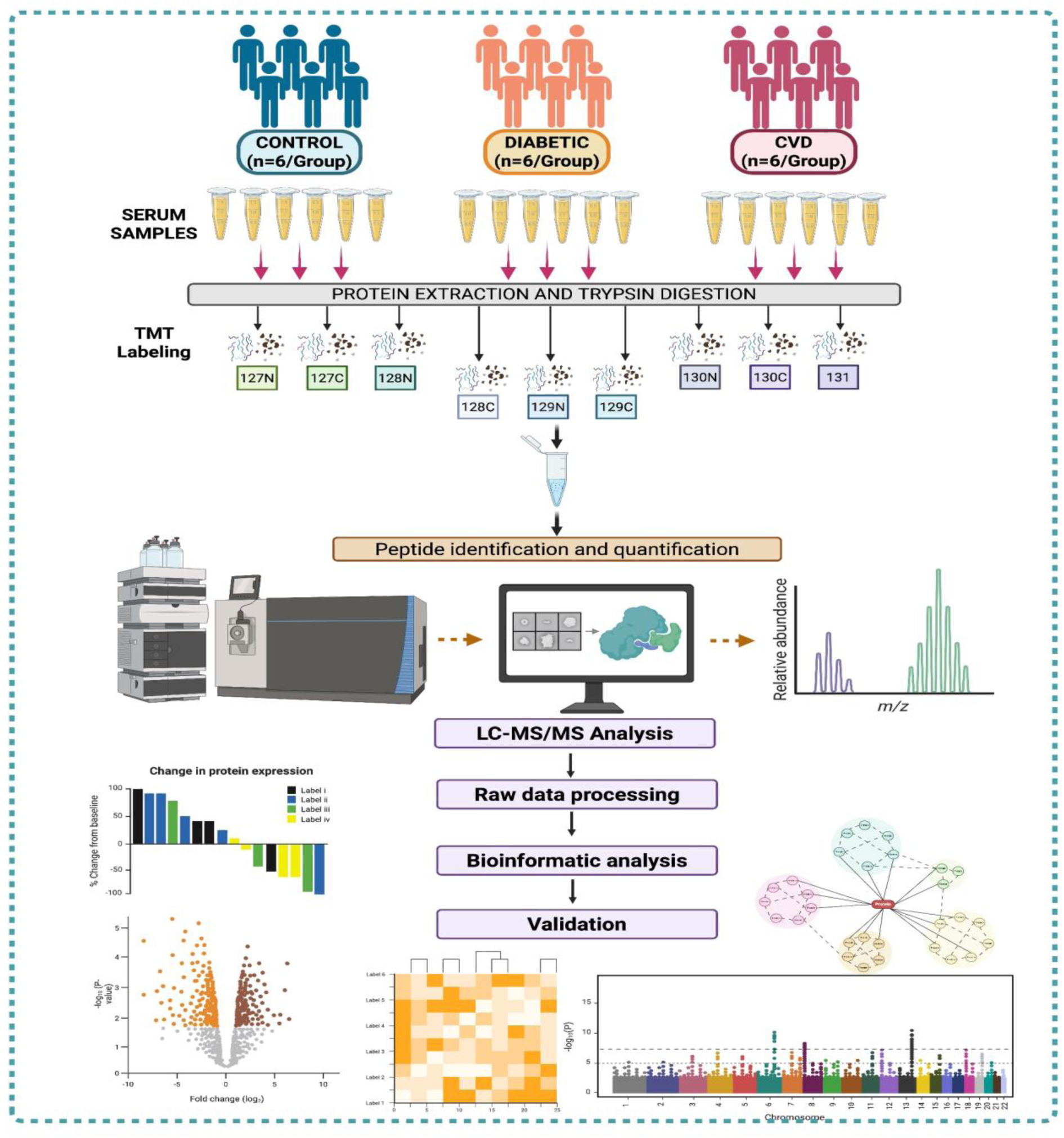
Experimental workflow for TMT-based quantitative serum proteomics from Control, Diabetic, and CVD cohorts. Serum samples from Control, Diabetic, and CVD groups (n=6 each) were processed for protein extraction, trypsin digestion, and TMT labeling (127N–131). Labeled peptides were pooled and analyzed by high-resolution LC- MS/MS for peptide identification and quantification. Raw spectra were processed for normalization and statistical analysis, followed by bioinformatic assessments including volcano plots, heatmaps, and network enrichment to identify disease-associated proteomic changes.

### LC-MS/MS Method and Data Searching

LC-MS/MS analysis was performed on a Q Exactive Plus mass spectrometer (Thermo Fisher Scientific, San Jose, CA) equipped with an EASY-Spray nanoESI source [36, 37]. Peptides (300ng) were eluted from an Acclaim Pepmap 100 trap column (75 µm ID × 2 cm, Thermo Scientific) onto an EASY-Spray PepMap Neo UHPLC analytical column (75 µm ID × 50 cm, Thermo Scientific using a 3-25% gradient of solvent B (acetonitrile, 0.1% formic acid) over 150 min, 25-50% solvent B over 20 min, 50-70% of solvent B over 10 min, 70-95% B over 10 min then a hold of solvent 95% B for 20 min, and finally a return to 3% solvent B for 10 min. Solvent A consisted of water and 0.1% formic acid. Flow rates were 300 nL/min using a Vanquish Neo UHPLC System (Thermo Fisher Scientific, San Jose, CA). Data dependent scanning was performed by the Xcalibur v 4.7.69.37 software [38] using a survey scan at 70,000 resolution scanning mass/charge (m/z) 375-1400 at an automatic gain control (AGC) target of 3e6 and a maximum injection time (IT) of 50 msec, followed by higher-energy collisional dissociation (HCD) tandem mass spectrometry (MS/MS) at 32 nce (normalized collision energy), of the 11 most intense ions at a resolution of 35,000, an isolation width of 1.2 m/z, an AGC of 1e5 and a maximum IT of 100 msec. Dynamic exclusion was set to place any selected m/z on an exclusion list for 30 seconds after a single MS/MS. Ions of charge state +1, 7, 8, >8, unassigned, and isotopes were excluded from MS/MS.

### Protein identification

MS and MS/MS data were searched against the amino acid sequence of the human database and a common contaminant protein database (e.g., trypsin, keratins; obtained at ftp://ftp.thegpm.org/fasta/cRAP) using Thermo Proteome Discoverer 3.1.1.93 [39] (Thermo Fisher Scientific). MS/MS spectra match considered fully tryptic peptides with up to 2 missed cleavage sites. Variable modifications considered were methylthio on Cys (+45.988Da) and met oxidation (+15.995Da). Proteins were identified at 95% confidence with XCorr score cut-offs as determined by a reversed database search.

### Quantitative data analysis

The protein and peptide identification results were further analyzed with Scaffold Q+S v 5.1.2 [40] (Proteome Software Inc., Portland OR), a program that relies on various search engine results (i.e.: Sequest, XTandem, MASCOT) and which uses Bayesian statistics to reliably identify more spectra. Protein identifications were accepted that passed a minimum of two peptides identified at 95% protein and peptide confidence levels. Quantitative values were generated using the Proteome Scaffold Q module. Acquired intensities in the experiment were globally normalized across all acquisition runs. Individual quantitative samples were normalized within each acquisition run. Intensities for each peptide identification were normalized within the assigned protein. The reference channels were normalized to produce a 1:1 fold change.

### Bioinformatics and Statistical Analysis

Processed MS spectral data were analyzed using bioinformatics tools in R Studio [41] and visualized as volcano plots, hierarchical clustering heatmaps, and protein–protein interaction networks to facilitate interpretation [42]. Statistical analyses were performed in GraphPad Prism 10.2.2 [43]. Comparisons between two groups were assessed using Student’s *t*-test, while Tukey’s multiple comparisons test was applied for analyses involving three or more groups. Statistical significance was defined as *p* < 0.05, \**p* < 0.01, and *p* < 0.001.

## Results

### Characteristics of the Study Population

Baseline biochemical parameters are summarized in (**Fig 2A)**. Both diabetic and CAD groups exhibited significantly elevated fasting blood glucose and HbA1c levels compared to controls (p < 0.001), confirming hyperglycemic status. Total cholesterol (TC) and low-density lipoprotein cholesterol (LDL-C) levels were lower in the disease groups, reflecting ongoing lipid-lowering treatment or altered lipid metabolism. High-density lipoprotein cholesterol (HDL-C) was markedly decreased in both diabetic (35.75 ± 8.31 mg/dL) and CAD (33 ± 6 mg/dL) patients compared to controls (49.14 ± 6.40 mg/dL), indicating impaired reverse cholesterol transport. Triglyceride (TG) levels were moderately elevated in diabetic (171.25 ± 53.47 mg/dL) and CAD (191.6 ± 165.5 mg/dL) groups relative to controls (156 ± 41.86 mg/dL). The TC/HDL ratio, a marker of atherogenic risk, was significantly higher in both disease cohorts (4.27 ± 0.81 in diabetic; 4.5 ± 1.8 in CAD) compared to controls (3.62 ± 0.76), further supporting an atherogenic lipid profile associated with cardiometabolic disease.

**Figure 2.**
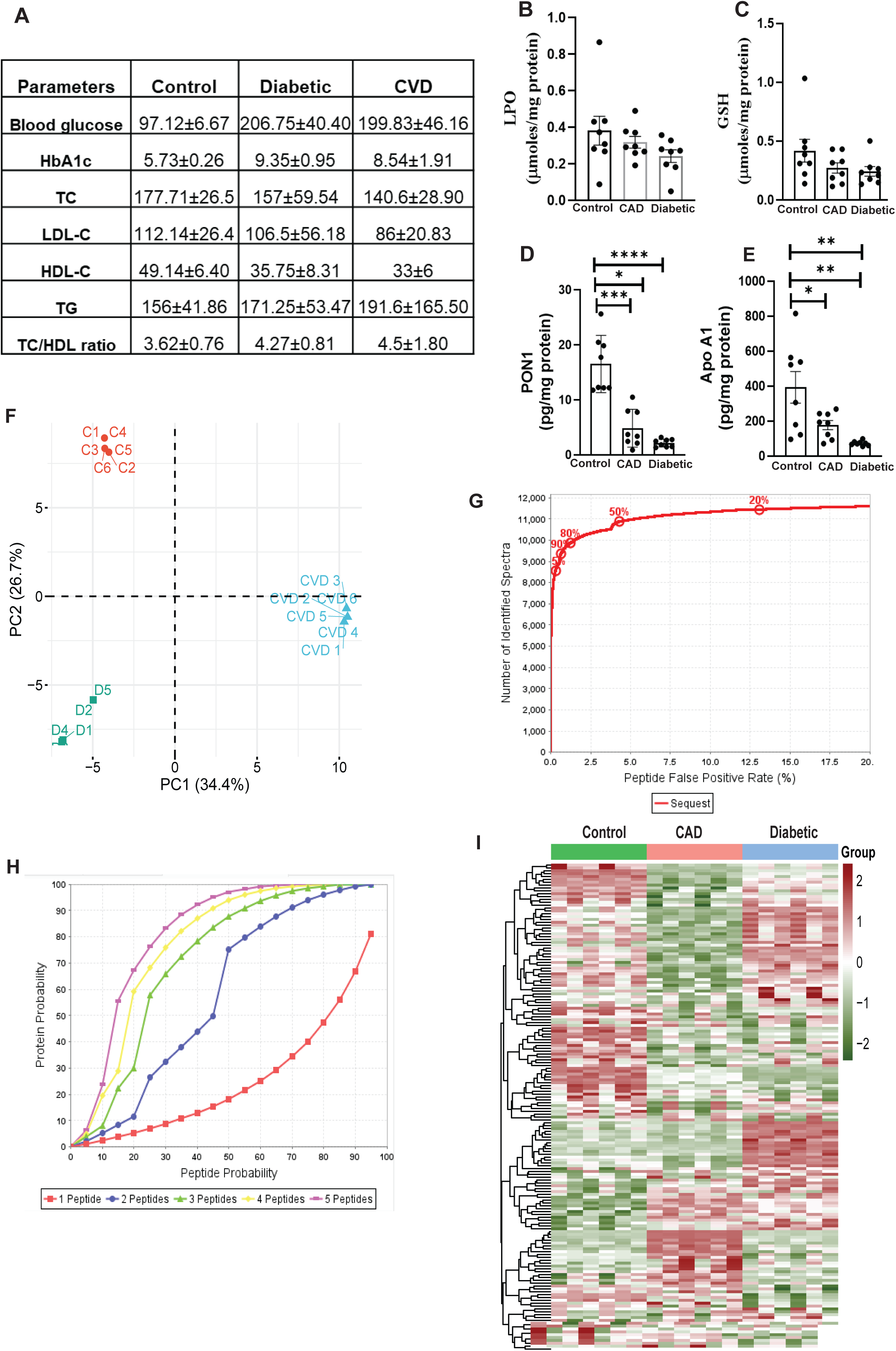
Clinical parameters and proteomic characterization of HDL dysfunctionality in Control, CAD and Diabetic subjects. (A) Clinical and biochemical characteristics of study participants. Table summarizes fasting blood glucose, HbA1c, total cholesterol (TC), LDL-C, HDL-C, triglycerides (TG), and TC/HDL ratio across Control, Diabetic, and CVD cohorts. Data expressed as mean ± SD. (B) Lipid Peroxidation (LPO) levels measured in isolated serum fractions from Control, CAD, and Diabetic groups. Increased LPO reflects heightened oxidative modifications in diseased cohorts. (C) GSH quantification in serum across the three groups, demonstrating lower GSH content in CAD and Diabetic subjects. (D) PON1 enzymatic activity in HDL fractions. Both CAD and Diabetic HDL displayed significantly suppressed paraoxonase activity relative to Control, consistent with dysfunctional antioxidant capacity. (E) ApoA-I abundance (relative ratio) derived from LC-MS/MS–based quantitation showing altered ApoA-I levels in CAD and Diabetic subjects. (F) Principal Component Analysis (PCA) of serum proteome depicting clear clustering separation between Control (green), CAD (red), and Diabetic (blue) groups along PC1 (34.4%) and PC2 (26.9%), indicating distinct proteomic signatures associated with disease status. (G) Peptide rank abundance distribution displaying cumulative contribution of peptide intensities. Curve highlights the top-abundant peptides driving group-wise proteomic 18 divergence. (H) Posterior error probability (PEP) curves for protein identification confidence across groups. Distribution shifts indicate differences in proteome complexity and detection depth among sample types. (I) Hierarchical clustering heatmap of significantly altered HDL- associated proteins across Control, CAD, and Diabetic cohorts. Rows represent proteins and columns represent individuals; color gradient (green to red) denotes relative abundance (downregulated to upregulated). Distinct clusters (Groups 1–3) highlight disease-specific proteomic remodeling.

### Serum Oxidative Stress and Lipid Metabolism Markers

To evaluate the redox–lipid imbalance underlying cardiovascular pathology, we examined serum biomarkers of oxidative stress and lipid metabolism across CAD, Diabetic, and healthy control cohorts **(Fig.2B-E)**. LPO levels were elevated in both CAD and Diabetic groups compared to controls, reflecting increased oxidative damage in disease states. GSH levels were significantly decreased in CAD and Diabetic patients, indicating compromised antioxidant defense. PON1 levels were markedly reduced in CAD and Diabetic groups, suggesting impaired HDL-associated antioxidant activity. Apo A1 levels were significantly lower in disease groups and reduction may contribute to dysfunctional lipid transport and heightened inflammatory response. Together, these findings underscore a pattern of oxidative stress and impaired lipoprotein metabolism in CAD and Diabetic patients.

### Distinct proteomic signatures among Control, CAD and Diabetic groups

Principal Component Analysis (PCA) of TMT data distinctly separates three groups- Control, CAD and Diabetic with first principal component (PC1) accounted for 34.4% of the total variance and the second principal component (PC2) accounted for 26.7% **(Fig.2F)**. Peptide-spectrum match (PSM) validation using the Sequest search algorithm in Scaffold demonstrated a strong correlation between the number of identified spectra and the peptide false positive rate (FPR) **(Fig.2G)**. The cumulative identification curve reached a plateau, indicating high identification confidence at low FPR thresholds. At a stringent 1% FPR, approximately 9,000 spectra were confidently identified, representing over 80% of all true-positive matches. Protein probability analysis demonstrated a strong correlation between peptide probability and the number of peptides assigned per protein **(Fig.2H)**. As expected, proteins identified with multiple unique peptides exhibited markedly higher confidence scores. Hierarchical clustering **(Fig.2I)** reveals distinct expression patterns among 3 groups, suggesting that the protein expression profiles are pathology-specific. Notably, samples within each group cluster closely together, indicating intra-group similarity and inter-group divergence. Together, these data indicate that changes in proteomic signatures are distinct in Diabetic vs control, CAD vs control and Diabetic vs CAD groups.

### Pathway enrichment analysis reveals disease-specific redox–lipid–proteostasis networks

Pathway enrichment in the control group **(Fig.3A)** demonstrated enrichment in cholesterol metabolism (APOE, APOC1, APOB, LCAT), ER protein processing (LBP, CPB2), and vascular smooth muscle contraction. Additional enriched pathways included PI3K-Akt, NF-κB, HIF-1, TGF-β, and glucagon signaling. These findings indicate stable metabolic and signaling networks in healthy individuals. CAD subject’s pathway enrichment **(Fig.3B)** revealed broad changes in cholesterol metabolism (LCAT, C8A, C9, APOB), ER protein processing (CPB2, LBP, IGFBP3), and stress-related signaling (NF-κB, HIF-1, TGF-β). PI3K-Akt signaling along with the enrichment of AGT, TFRC, and KRT10 reflects proliferative and stress responses. Additional enrichment in PPAR signaling, ferroptosis, and cysteine/methionine metabolism suggests metabolic reprogramming and oxidative stress.

**Figure 3.**
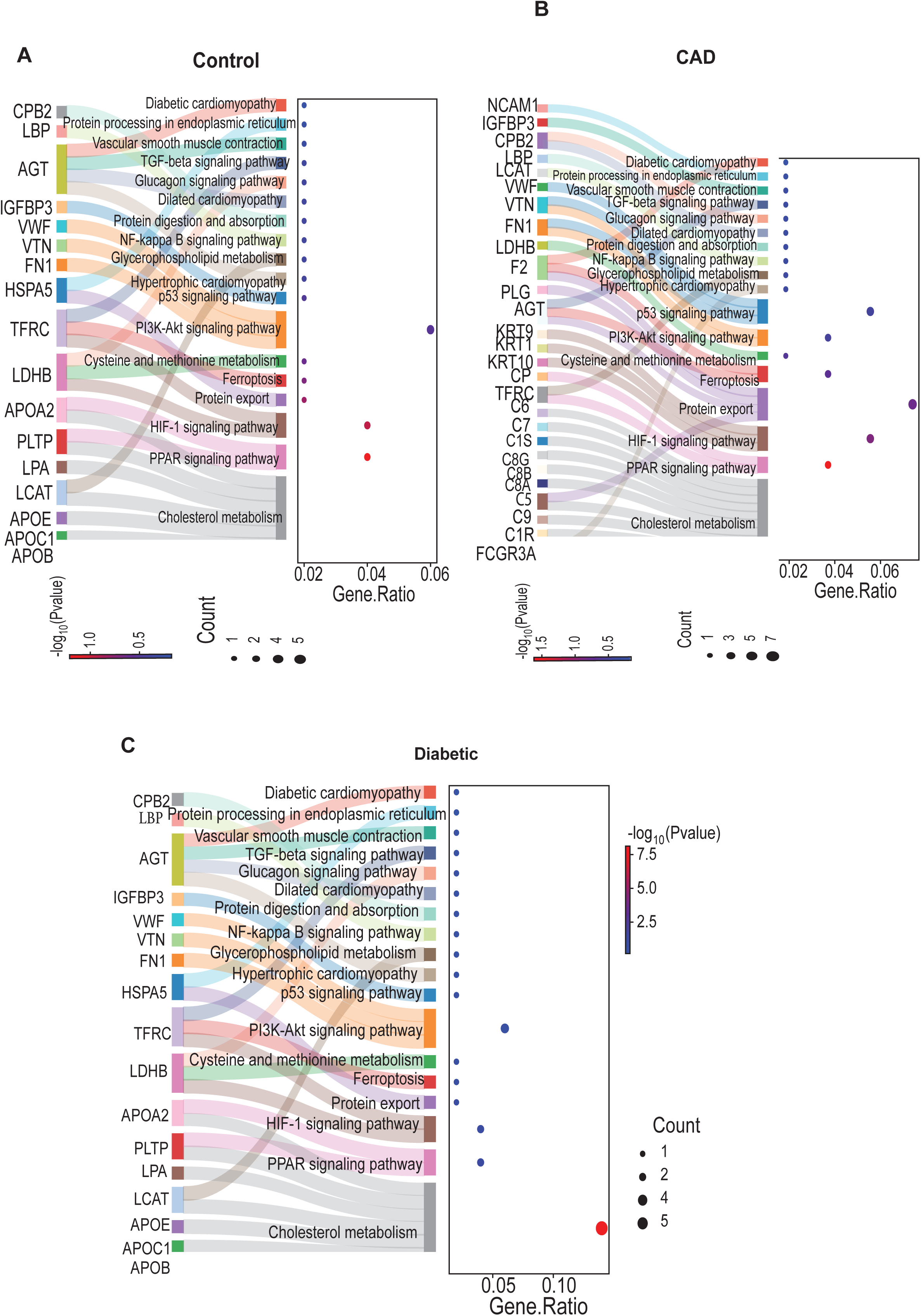
Pathway enrichment analysis serum proteins in Control, CAD, and Diabetic groups. (A–C) Sankey plots and dot plots showing KEGG pathway enrichment of differentially abundant serum proteins in Control (A), CAD (B), and Diabetic (C) groups. Key pathways represented include cholesterol metabolism, PPAR signaling, PI3K–Akt signaling, ferroptosis, ECM remodeling, inflammatory signaling (NF-κB, HIF-1), and cardiomyopathy-related pathways. Dot size indicates the number of proteins enriched per pathway, and color reflects –log10(p- value). These analyses highlight distinct pathway perturbations across clinical groups, reflecting disease-specific remodeling of the HDL proteome.

Diabetes-associated enrichment **(Fig.3C)** implicated genes such as APOE, FN1, HSPA5, and APOB across multiple pathways, underscoring their uniqueness from CAD group. PI3K-Akt signaling was most significantly enriched (gene ratio 0.24, –log10P = 8.5), along with cholesterol metabolism and ER protein processing, consistent with metabolic dysregulation. Enrichment of Diabetic and hypertrophic cardiomyopathy, TGF-β, and NF-κB pathways reflect the cardiovascular and inflammatory burden in diabetes group. Together, these analyses show that control, CAD, and diabetes segregate into distinct redox–lipid–proteostasis states.

### Differential expression of Apolipoproteins among the Diabetic and CAD patients

To delineate disease-specific alterations in circulating apolipoproteins, we compared their relative expression patterns across control, diabetic, and CAD cohorts (**Fig. 4A**). The analysis revealed distinct group-specific trends in apolipoprotein abundance. Diabetic plasma exhibited a selective upregulation of APOC1, APOM, and APOD, reflecting activation of stress-responsive and lipid-remodeling apolipoproteins. In contrast, the CAD group displayed a broader dysregulation pattern, characterized by increased expression of APOC1, APOM, and APOD, together with variable changes in APOB, indicating a shift toward an inflammatory and atherogenic lipid profile. Core HDL-associated apolipoproteins- APOA1, APOA2, and APOA4, remained relatively stable across groups, suggesting preservation of basal lipid transport capacity. Overall, diabetic samples showed targeted modulation of specific apolipoproteins linked to metabolic stress, whereas CAD samples demonstrated more extensive alterations consistent with combined oxidative, inflammatory, and lipid-handling dysfunction.

**Figure 4.**
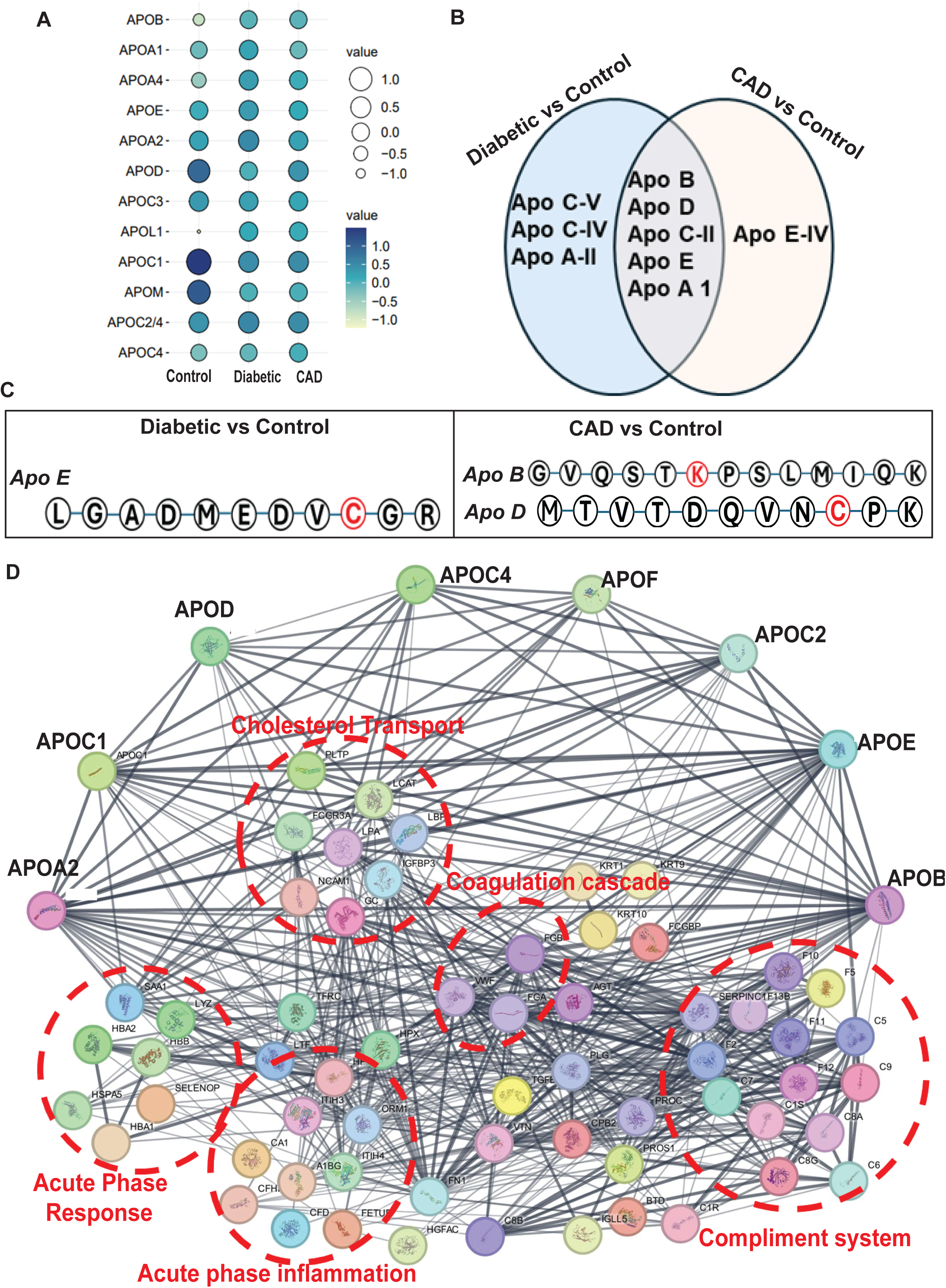
Apolipoprotein remodeling, post-translational modifications, and protein– protein interaction networks reveal disease-specific apolipoprotein proteoform signatures in Diabetic and CAD subjects. (A) Dot-plot visualization of differentially abundant apolipoproteins across Control, Diabetic, and CAD groups. (B) Venn diagram illustrating shared and unique apolipoproteins altered in Diabetic vs Control and CAD vs Control groups. (C) Post-translationally modified peptide sequences identified in Diabetic and CAD subjects. (D) Protein– protein interaction (PPI) network of differentially abundant apolipoproteins across groups.

Venn diagram **(Fig.4B)** demonstrates that in the Diabetic vs control group, three apolipoproteins were uniquely altered- Apo C-V, Apo C-IV, and Apo A-II, suggesting these may be specifically associated with Diabetic metabolic dysregulation. In contrast, the CAD vs Control group showed a unique alteration in Apo E-IV, indicating a potential role of Apo E-IV in cardiovascular pathology independent of diabetes. Importantly, five apolipoproteins Apo B, Apo D, Apo C-II, Apo E, and Apo A-I were found to be commonly dysregulated in both Diabetic and CAD patients. This overlap suggests a shared proteomic signature linked to underlying atherosclerotic or inflammatory processes common to both conditions.

### Identification of potential amino acid modifications in apolipoproteins in Diabetic vs. CAD patients

To investigate the molecular alterations in apolipoproteins associated with diabetes and CAD, we analyzed post-translational modifications (PTMs) in amino acid residues **(Fig.4C)**. In Diabetic samples, the cysteine residue within *Apo E* showed oxidative modification. This alteration likely disrupts Apo E’s structural stability and its lipid-binding affinity, contributing to impaired cholesterol efflux and HDL dysfunction observed in diabetes. In the CAD group, oxidation was detected at lysine residues in *Apo B* and cysteine residues in *Apo D*, suggesting distinct redox-driven modifications of both lipid-transport and stress-responsive apolipoproteins. Oxidized lysine in *Apo B* is typically associated with increased susceptibility to LDL aggregation and atherogenicity, while cysteine oxidation in *Apo D* may impair its antioxidant roles. Together, these site-specific oxidative modifications reveal disease-dependent redox remodeling of apolipoproteins, linking systemic oxidative stress to functional impairment of lipid transport pathways in both diabetes and CAD.

### Post-translationally modified apolipoproteins as central nodes in linking cholesterol handling to vascular inflammation

To identify apolipoprotein involved pathways and those potentially disturbed by their PTMs, we performed protein–protein interaction (PPI) network analysis **(Fig.4D)**. Within this network, APOB and APOE emerged as hub nodes acting as bridging points to subnetworks -coagulation cascade, complement system, and acute-phase inflammation. The cholesterol transport mediators- APOC1/2/4, APOF, CETP, LCAT etc. showed strong interconnectivity. Notably, this lipid-handling cluster displayed multiple edges toward coagulation factors (F2, F5, F10, fibrinogen chains). Similarly, extensive wiring with complement proteins (C3, C5–C9, CFH, CFB) highlighted a role for lipoproteins in innate immune surveillance. While acute-phase proteins (CRP, SAA1/2, haptoglobin) formed connections that reflect HDL remodeling during systemic inflammation. This cross-module connectivity reveals that lipid metabolism, thrombosis, complement activation, and inflammatory responses are not independent processes but are coordinately regulated through apolipoprotein hubs. Given this network connection, PTMs on any apolipoproteins alter cholesterol handling, coagulation, and immune pathways leading to metabolic imbalance and vascular pathology.

## Discussion

In this study, we provide the first integrated, redox-informed serum proteomic comparison distinguishing CAD alone from CAD with diabetic comorbidity. Our findings reveal that while both conditions share a background of oxidative stress and lipoprotein dysregulation, diabetes imposes an additional layer of metabolic and redox remodeling that results in distinct apolipoprotein signatures and oxidative post-translational modifications (PTMs). These results extend current understanding of how diabetes amplifies CAD risk and offer molecular insight into why diabetic coronary disease presents with accelerated atherosclerosis, calcification, and greater clinical complexity.

Previous proteomic studies have reported elevated inflammatory mediators, complement activation, and perturbations in lipid transport pathways in CAD and diabetes independently [44–46]. However, most of these datasets examined each disease in isolation. Our TMT-based quantitative proteomics reveals that when diabetes coexists with CAD, the serum proteome is reshaped in a way that is not simply additive but reflects synergistic metabolic and redox stress. Consistent with earlier work highlighting enhanced carbonyl stress and ER dysfunction in diabetes [47, 48], our pathway enrichment analysis identifies pronounced activation of ER protein processing, PI3K-Akt/mTOR, and TGF-β signaling in the diabetic cohort pathway, while less enriched in CAD alone. These diabetes-dominant pathways likely contribute to maladaptive lipoprotein remodeling, endothelial injury, and impaired cholesterol efflux that accelerate atherogenesis in diabetic patients.

A novel finding of this work is the identification of disease-specific oxidative PTMs on circulating apolipoproteins, providing mechanistic insight into how redox imbalance disrupts lipid-transport function. Previous studies have established that oxidative modification of apolipoproteins particularly ApoA-I and ApoB impairs HDL-mediated cholesterol efflux and enhances LDL aggregation [49, 50]. We extend this knowledge by mapping site-specific cysteine oxidation in ApoE uniquely in diabetic subjects, and lysine oxidation in ApoB and cysteine oxidation in ApoD uniquely in CAD subjects. These PTMs likely alter lipoprotein conformation, receptor affinity, and interactions with cellular scavenger receptors, thus providing a molecular explanation for the exaggerated lipoprotein dysfunction characteristic of cardiometabolic disease. The observation that ApoD oxidation is CAD-specific is a novel observation [51], as ApoD has emerging roles in lipid peroxidation buffering and cellular stress protection, its modification may compromise antioxidant function and contribute to plaque vulnerability.

Our PPI network analysis further shows that apolipoproteins modified in disease ApoB, ApoD, ApoE, and ApoC isoforms serve as hub proteins linking lipid transport modules to the complement system, coagulation cascade, and acute-phase inflammation. This observation is consistent with recent systems biology frameworks demonstrating that lipoproteins act not only as lipid carriers but also as platforms integrating immune and coagulation signals during cardiometabolic stress [52, 53]. The convergence of these subnetworks in our diabetic and CAD proteomes suggests that PTMs in apolipoproteins may propagate broader disturbances in vascular inflammation, thrombosis, and immune activation. Such crosstalk may partially explain why diabetic CAD patients experience higher rates of microvascular dysfunction and plaque vulnerability

Importantly, our dataset identifies multiple apolipoproteins ApoC-V, ApoC-IV, and ApoA-II uniquely dysregulated in diabetic serum, implicating these proteins in diabetes-specific lipid remodeling. ApoC isoforms regulate lipoprotein lipase (LPL) activity and triglyceride clearance [54], their selective dysregulation may drive hypertriglyceridemia and small, dense LDL formation typical of diabetic dyslipidemia. In contrast, CAD-specific alteration of ApoE-IV suggests an isoform-dependent signature of cardiovascular pathology independent of metabolic disease. The five apolipoproteins dysregulated in both CAD and diabetes (ApoB, ApoD, ApoC-II, ApoE, and ApoA-I) likely represent shared atherogenic indices, while the unique signatures highlight disease-specific mechanisms that may support precision biomarker development.

From a translational perspective, our integrated proteomic and redox analysis emphasizes the need to consider HDL functionality and apolipoprotein oxidation rather than HDL-C levels alone when assessing cardiovascular risk. Although both CAD and diabetic patients exhibited reduced HDL-C, our findings indicate that oxidative remodeling of apolipoproteins rather than concentration may be the primary determinant of HDL dysfunction in cardiometabolic disease. This aligns with growing clinical evidence that HDL function, not HDL quantity, predicts cardiovascular events.

## Author Approval

All authors have approved the final version of the manuscript.

## Competing Interests

No conflict of interest to disclose.

## Authors Contributions

RNS, KP, and SS contributed to the conceptualization and overall study design. SS and AR conducted the data analysis and prepared the initial manuscript draft. NZ and KP performed the TMT labeling and LC–MS experiments. AR, GP, and SM contributed to sample collection, processing, and isolation. AKC provided critical review and revision of the manuscript. SS also provided mentorship and guidance to the team throughout the study.

## Data Availability

All data generated in this study are available from the corresponding author upon reasonable request. The proteomics datasets will be deposited in the PRIDE database and made publicly accessible upon acceptance of the manuscript.

## Notes

### Competing Interest Statement

The authors have declared no competing interest.

### Funding Statement

This study did not receive any funding.

### Author Declarations

The study was conducted in accordance with the Declaration of Helsinki and approved by the institutional ethics committees of all participating institutions (University of Madras: UM/IHEC/F.RM/2021-XIV; Global Gleneagles Hospitals: BMHR/2024/0076; Sri Ramachandra Medical College (SRMC) : IEC-NI/21/APR/78/85).

